# Association between urbanicity and depressive symptoms among Chinese middle-aged and older adults

**DOI:** 10.1101/2024.09.02.24312930

**Authors:** Yanhua Chen, Peicheng Wang, Qiaoyuan He, Jiming Zhu, Mika Kivimäki, Gill Livingston, Andrew Sommerlad

**Author notes:** Corresponding Author: Yanhua Chen. Andrew Sommerlad.

## Abstract

**Background:** Depression is a pressing public health issue and may be affected by multifaceted urban living, yet the specific urbanicity elements associated is unclear. Using a multidimensional urbanicity scale, we explored the association between urbanicity and its components with the risk of depressive symptoms.

**Methods:** This study used data from four waves of the China Health and Retirement Longitudinal Study, including 12,515 participants aged ≥45 years at baseline in 2011 in 450 rural and urban communities, and 8,766 with 7 years of follow-up. Multilevel logistics regression and Cox proportional hazards regression models examined the cross-sectional and longitudinal associations between urbanicity and depressive symptoms.

**Results:** Living in areas with the highest tertile of urbanicity was associated with a 61% lower risk of depressive symptoms cross-sectionally (odds ratio (OR): 0.39, 95% confidence interval (CI): 0.30-0.50) and 33% lower risk longitudinally (hazard ratio (HR): 0.67, 95% CI: 0.58-0.77) compared to those living in areas with the lowest tertile of urbanicity. Among components, higher population density (OR: 0.92, 95% CI: 0.87-0.97), better education (OR: 0.94, 95% CI: 0.89-0.99), transportation (OR: 0.95, 95% CI: 0.92-0.98), sanitation (OR: 0.96, 95% CI: 0.93-0.98) was associated with lower odds of depressive symptoms, while greater population educational and socioeconomic diversity (OR, 1.08; 95% CI, 1.03-1.13) had opposite effect. Better economic conditions (HR: 0.94, 95% CI: 0.90-0.98) and availability of social services (HR, 0.96; 95% CI, 0.93-0.99) were longitudinally associated with reduced risk of developing depressive symptoms during 7 years of follow-up. Additionally, differences in associated components were found between urban and rural residents and between midlife and older adults.

**Conclusions:** Our findings underscore the complex links of urban living with depressive symptoms among middle-aged and older adults, highlighting the need to consider a multidimensional urbanicity perspective to understand the urbanicity-mental health nexus. Tailored urban planning policies should consider the associated urbanicity components, along with temporal effectiveness, urban-rural disparities, and age group differences.

## Introduction

Depression in midlife and older adults is an increasing public health problem^1,2^, as it is associated with emotional suffering, reduced physical, cognitive and social functioning, increased medical costs, risk of suicide, and all-cause and cardiovascular mortality^3,4,5,6^. Effective interventions preventing depression in this vulnerable group are of public health importance. Urban development significantly influences human health^7,8,9^ and urban environmental exposures are recognized as potential risk factors for mental health disorders^10^. A substantial body of literature indicates that mental health problems are more prevalent in urban areas compared to rural ones^11^, with much of this research centered in high income countries (HICs)^12,13,14,15^. A meta-analysis found that, in HICs, living in urban areas was associated with a greater likelihood of depressive disorder compared to rural areas but this association was not observed in low and middle-income countries (LMICs), nor in samples of older adults or children/adolescents^16^.

Urbanicity refers to the impact of living in urban areas ^17^ and urban-living has complex positive and negative interactions with health^18^. For example, urban areas attract residents due to better access to healthcare, education and work opportunities; while urban dwellers often encounter specific challenges such as exposure to pollutants, limited green spaces and crowding^19^. Given urban living’s multifaceted effects, relying as previous studies have done on simple urban/rural dichotomy^20,21,22^, or single item proxy, like population density^23^ or urbanization rates^24^, fails to capture the nuanced differences emerging within urban and rural communities^25,26^. Instead, a multidimensional urbanicity scale would offer a more comprehensive and informative measure of urbanicity variations^27^, and can also elucidate which urban living elements most significantly affect health, and therefore guide preventative approaches^28,29,30^. Recent studies reporting that urban areas may increase or decrease risk of depressive symptoms among older adults^31,32^ have predominantly used typical urban-rural dichotomy or single-item continuum (i.e., population density or land use)^33,32^. There remains a paucity of evidence using a multidimensional urbanicity scale to explore the association of urbanicity and its components with depressive symptoms, and this understanding may inform future targeted public health interventions for middle-aged and older adults in urban and rural areas.

We therefore aim to examine the relationship between urbanicity and depressive symptoms. Our specific objectives are

1. To examine the cross-sectional and longitudinal associations between urbanicity and significant depressive symptoms, given the cross-sectional data of might not accurately distinguish the longitudinal cumulative effects of urbanicity on health^34^.
2. To examine how twelve urbanicity components influence their significant depressive symptoms, with basis of a multicomponent urbanicity index created for and widely used in China’s context^25^.
3. Considering the urban-rural disparities in the China and the variation in depressive symptoms across age groups^35,36^, we will assess if any associations vary based on urban or rural residency, and by midlife and older adulthood.

## Methods

### Study population

We obtained data from baseline (2011) and three follow-up surveys (201, 2015, 2018) of the China Health and Retirement Longitudinal Study (CHARLS, accessible at http://charls.pku.edu.cn/). CHARLS is a nationally-representative survey focuses on individuals aged 45 and above in China^37^. Using a stratified multi-stage “probability proportional to size” random sampling method, CHARLS sampled residents from 28 provinces, with a total of 17,708 individuals were survey at the baseline. CHARLS encompasses a wide range of information at both personal- and community-level. Since CHARLS only conducted a community survey in its baseline year of 2011, we also supplemented city-level administrative data from the China City Statistical Yearbooks and China Statistical Yearbook for Regional Economy in 2011 to the study to provide information on the economic situation and medical resources. We then linked the individual- and area-level datasets by CHARLS primary sampling unit (PSU) code, and our analysis ultimately included 12,515 individuals in 2011 and 8,766 follow-up participants from 2013 to 2018 who were without significant depressive symptoms at baseline. Participants lived in 450 communities nested within 126 cities. A flowchart of participants is illustrated in Figure S1.

Ethical approval was obtained from Peking University’s Ethical Review Committee (IRB000001052-11015) and all participants gave written informed consent before participating in the study.

### Measurements

#### Significant depressive symptoms

Mental health was assessed in all four waves using the 10-item version of the Center for Epidemiologic Studies Depression (CES-D) Scale^37^, which has a score range of 0 to 30. The CES-D has been widely used in mental health research and as a tool for detecting depressive symptoms in older adults. We categorized participants as having no significant depressive symptoms if CES-D <12 and significant depressive symptoms if ≥ 12, and this cutoff point has been proven reliable and valid in previous studies^38,39^. We also used CES-D scores as a continuous variable in secondary analyses; higher scores of CES-D indicate poorer mental health status.

#### Urbanicity

Urbanicity was assessed at baseline in 2011 using a multidimensional urbanicity index developed using CHARLS community survey and Statistical Yearbooks by taking into account the population characteristics, data availability, and components of urbanicity based on a priori hypotheses and previous research^25,40,41^. The index comprised 12 urbanicity components: population density, economic conditions, market/commercial development, medical facilities availability and accessibility, transportation availability and quality, housing facilities quality, availability and use of communication and technologies, provision of sanitation services and facilities, educational attainment, social service availability, community facilities availability, and population educational and socio-economic diversity. Each component was synthesized by 1 to 5 factors. For detailed descriptions of each urbanicity component and its source, see Table S1. Each component was scaled from 0 to 10 and then added together for a possible range of 0-120. Higher scores indicated a greater degree of urbanicity. We further classified urbanicity levels into low, moderate, and high based on tertiles of urbanicity scores. Multiple imputation was used to address the missing data in urbanicity components (see Table S2).

#### Covariates

Individual information on sociodemographic characteristics, lifestyle behaviors and health-related factors were collected at baseline using questionnaires. Covariates included age (continuous variable), sex (male or female), education (illiterate meaning limited or no access to formal education, primary school, middle school, high school/vocational high school, or college and above), marital status (married and partnered status or otherwise), residence (urban or rural), socioeconomic status (quintiles of per-capita household consumption expenditure: quantile 1, <2,552 Chinese Yuan (CNY) per annum; quantile 2, 2,553-4,132 CNY; quantile 3, 4,133-6,255 CNY; quantile 4, 6,256-10,268 CNY, quantile 5, (>=10,269 CNY), living status (alone or with others), smoking (currently smoking or ex/never smoked), drinking (currently drinking or ex/never drinking), and chronic diseases (having one or more chronic diseases or none).

### Statistical analysis

For descriptive statistics, we calculated the prevalence and percentages of respondents at baseline, categorized by all covariates and whether they had significant depressive symptoms. Continuous variables, such as age, were further summarized using mean and standard deviation, and per-capita household consumption expenditure using median and interquartile range. In the main analyses, we applied multilevel logistic regression for cross-sectional data as baseline year and Cox proportional hazards regression for 7-year follow-up data, respectively, to evaluate the association of urbanicity with depressive symptoms.

In the cross-sectional analysis, we estimated multilevel mixed-effects models, incorporating a random intercept for each community to account for the nesting of individuals within communities. The intra-class correlation coefficient (ICC) at the community level in the null model was 0.146, suggesting that a multilevel approach is advisable, with 14.6% of depressive symptoms potentially attributable to community factors. Odds ratios (OR) with corresponding 95% confidence intervals (CIs) in multilevel logistic regression models to demonstrate the effects of cross-sectional associations. In longitudinal analysis, we employed Cox proportional hazards regression to examine the longitudinal association between urbanicity at baseline and the subsequent incidence of significant depressive symptoms, with hazard ratios (HR) with 95% confidence intervals reported. Participants who either did not develop significant depressive symptoms during follow-up or were lost to follow-up were considered as censored data in this study. In both cross-sectional and longitudinal analyses, we performed the main model with crude model and multivariable-adjusted Subgroup analyses were conducted according to type of residence (i.e., urban or rural) and age group (i.e., middle-aged or older) models. In all analyses, age, sex, education, marital status, socioeconomic status, living status, smoking, drinking and chronic disease status were added in the full-adjusted models. All analyses were conducted using Stata (version 17.0; StataCorp) and the statistical significance was determined at the p < 0.05 level.

#### Sensitivity analyses

We conducted a series of sensitivity analyses to assess the robustness of the associations between urbanicity and depressive symptoms. We repeated the main analyses using urbanicity scores ranging from 0 to 120, using cut-off of 10 to define significant depressive symptoms^42^ and excluding individuals whose urbanicity has missing data. We further estimated the different effects of 12 urbanicity components and stratified the associations by urban/rural residence. A multicollinearity analysis of urbanicity components was conducted using the condition indexes, indicating the absence of collinearity (see Table S3).

## Results

The characteristics of the 12,515 participants included at baseline are in Table 1. 3,004 participants (24.0%) exhibited significant depressive symptoms. 6,476 (51.8%) were female and 6,039 (48.3%) were male, and 3,307 (26.4%) were aged ≥65 years with a total mean 59.0 years (SD 9.5). The majority had an educational level below primary school (8,328 individual, 66.5%), lived in rural areas (7,640 individuals, 61.1%) and only 696 (5.6%) lived alone. Their median annual per-capita household consumption expenditure was 5,020.0 (IQR 6054.0) Chinese Yuan (CNY). Approximately one-third of the participants reported currently drinking alcohol (4,082 individuals, 32.6%) or smoking (3,843 individuals, 31.7%), and more than three-fifths (8,534, 68.2%) had chronic diseases. The distribution of the follow-up participants was similar (see table S4).

**Table 1.** Baseline demographic characteristics of the study participants.

| Variables: n (%) unless specified | Total (n = 12,515) |
| --- | --- |
| Age (years): mean (SD) | 59.0 (9.5) |
| 45-54 | 4,480 (35.8) |
| 55-64 | 4,738 (37.9) |
| 65-74 | 2,371 (19.0) |
| $\geq 75$ | 926 (7.4) |
| Sex |  |
| Male | 6,039 (48.3) |
| Female | 6,476 (51.8) |
| Education |  |
| Illiterate | 5,660 (45.2) |
| Primary school | 2,668 (21.3) |
| Middle school | 2,593 (20.7) |
| High school/vocational high school | 1,328 (10.6) |
| College and above | 266 (2.1) |
| Married status |  |
| Married and partnered | 10,487 (83.8) |
| Other | 2,028 (16.2) |
| Socioeconomic status (annual per-capita household consumption expenditure): median (IQR) | 5,020.0 (6054.0) |
| Quintile 1 (<2,552 CNY) | 4,379 (35.0) |
| Quintile 2 (2,553-4,132 CNY) | 3,184 (25.4) |
| Quintile 3 (4,133-6,255 CNY) | 2,320 (18.5) |
| Quintile 4 (6,256-10,268 CNY) | 1,696 (13.6) |
| Quintile 5 ($\geq 10,269$ CNY) | 936 (7.5) |
| Residency |  |
| Urban | 4,875 (38.9) |
| Rural | 7,640 (61.1) |
| Living alone |  |
| Yes | 696 (5.6) |
| No | 11,819 (94.4) |
| Current alcohol-drinker |  |
| Yes | 4,082 (32.6) |
| No | 8,433 (67.4) |
| Current smoker |  |
| Yes | 3,843 (31.7) |
| No | 8,672 (69.3) |
| Has one or more chronic disease |  |
| Yes | 8,534 (68.2) |
| No | 3,981 (31.8) |
| Significant depressive symptom (CES-D$\geq 12$) | |
| Yes | 3,004 (24.0) |
| No | 9,511 (76.0) |

Abbreviations: CES-D, Center for Epidemiological Studies Depression Scale; CNY, Chinese Yuan; IQR, Interquartile Range; SD, standard deviation.

Table 2 shows the results of cross-sectional and longitudinal associations between urbanicity and significant depressive symptoms. In cross-sectional analyses, after multivariable-adjustments (model 2), we found that living in areas within the moderate urbanicity was associated with a 27% lower odds of experiencing significant depressive symptoms (OR, 0.73; 95% CI, 0.61-0.88), compared to those in the low urbanicity area (lowest tertile). Living in the high urbanicity area was linked to a 61% lower risk (OR, 0.39; 95% CI, 0.30-0.50) compared with the low urbanicity. In the longitudinal analysis over a 7-year follow-up period (fully adjusted, Model 2), residing in a high urbanicity area was associated with a 33% reduction in the odds of developing significant depressive symptoms compared to those living in low urbanicity areas (HR: 0.67, 95% CI: 0.58-0.77); while living in a moderate urbanicity area showed no significant association (HR: 0.92, 95% CI: 0.84-1.01).

**Table 2.**
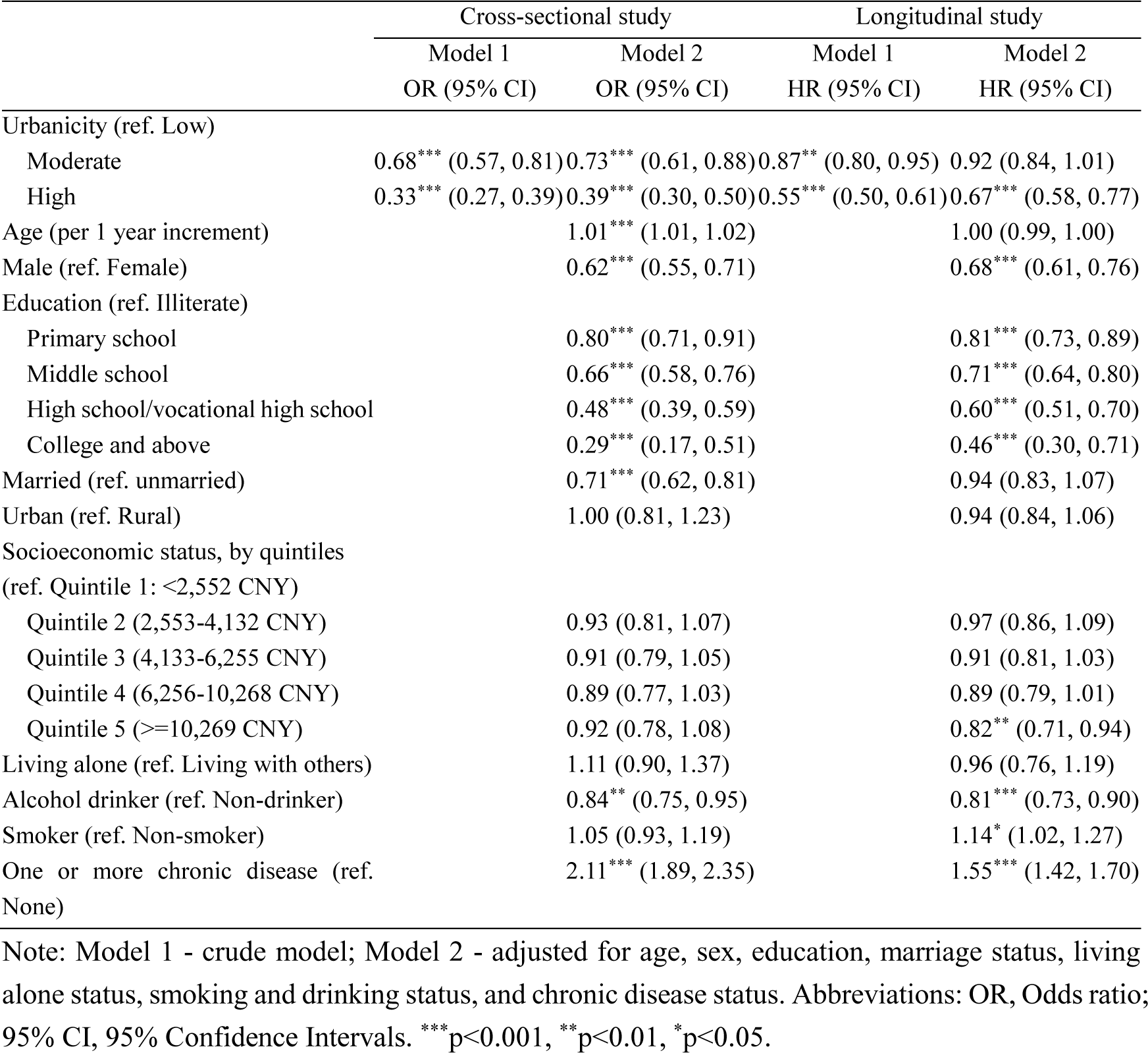
Cross-sectional and longitudinal associations between urbanicity and significant depressive symptoms.

|  | Cross-sectional study |  | Longitudinal study |  |
| --- | --- | --- | --- | --- |
|  | Model 1 | Model 2 | Model 1 | Model 2 |
|  | OR (95% CI) | OR (95% CI) | HR (95% CI) | HR (95% CI) |
| Urbanicity (ref. Low) |  |  |  |  |
| Moderate | 0.68*** (0.57, 0.81) | 0.73*** (0.61, 0.88) | 0.87** (0.80, 0.95) | 0.92 (0.84, 1.01) |
| High | 0.33*** (0.27, 0.39) | 0.39*** (0.30, 0.50) | 0.55*** (0.50, 0.61) | 0.67*** (0.58, 0.77) |
| Age (per 1 year increment) |  | 1.01*** (1.01, 1.02) |  | 1.00 (0.99, 1.00) |
| Male (ref. Female) |  | 0.62*** (0.55, 0.71) |  | 0.68*** (0.61, 0.76) |
| Education (ref. Illiterate) |  |  |  |  |
| Primary school |  | 0.80*** (0.71, 0.91) |  | 0.81*** (0.73, 0.89) |
| Middle school |  | 0.66*** (0.58, 0.76) |  | 0.71*** (0.64, 0.80) |
| High school/vocational high school |  | 0.48*** (0.39, 0.59) |  | 0.60*** (0.51, 0.70) |
| College and above |  | 0.29*** (0.17, 0.51) |  | 0.46*** (0.30, 0.71) |
| Married (ref. unmarried) |  | 0.71*** (0.62, 0.81) |  | 0.94 (0.83, 1.07) |
| Urban (ref. Rural) |  | 1.00 (0.81, 1.23) |  | 0.94 (0.84, 1.06) |
| Socioeconomic status, by quintiles<br>(ref. Quintile 1: <2,552 CNY) |  |  |  |  |
| Quintile 2 (2,553-4,132 CNY) |  | 0.93 (0.81, 1.07) |  | 0.97 (0.86, 1.09) |
| Quintile 3 (4,133-6,255 CNY) |  | 0.91 (0.79, 1.05) |  | 0.91 (0.81, 1.03) |
| Quintile 4 (6,256-10,268 CNY) |  | 0.89 (0.77, 1.03) |  | 0.89 (0.79, 1.01) |
| Quintile 5 (>=10,269 CNY) |  | 0.92 (0.78, 1.08) |  | 0.82** (0.71, 0.94) |
| Living alone (ref. Living with others) |  | 1.11 (0.90, 1.37) |  | 0.96 (0.76, 1.19) |
| Alcohol drinker (ref. Non-drinker) |  | 0.84** (0.75, 0.95) |  | 0.81*** (0.73, 0.90) |
| Smoker (ref. Non-smoker) |  | 1.05 (0.93, 1.19) |  | 1.14* (1.02, 1.27) |
| One or more chronic disease (ref. None) |  | 2.11*** (1.89, 2.35) |  | 1.55*** (1.42, 1.70) |

Note: Model 1 - crude model; Model 2 - adjusted for age, sex, education, marriage status, living alone status, smoking and drinking status, and chronic disease status. Abbreviations: OR, Odds ratio; 95% CI, 95% Confidence Intervals. \*\*\*p<0.001, \*\*p<0.01, \*p<0.05.

We conducted a series of sensitivity analyses. The results of the main analysis, showing a protective role of urbanicity on significant depressive symptoms in both cross-sectional and longitudinal associations, were consistent with the results of the sensitivity analysis using urbanicity score (Table S5), using different cut-off points for defining significant depressive symptoms (Table S6), using a pre-imputation dataset (Table S7). We further stratified the associations by type of residence and age group (Table S8-9). The protective effect of urbanicity was slightly lower among urban residents compared to rural residents. Among older adults, the estimated effect was similar with that of middle-aged adults in the cross-sectional analysis while not significant in the longitudinal analysis.

The results of the cross-sectional and longitudinal associations between urbanicity components and significant depressive symptoms are presented in Figure 1 and Table S10. Generally, among 12 urbanicity components, higher population density (OR, 0.92; 95% CI, 0.87-0.97), higher education (OR, 0.94; 95% CI, 0.89-0.99), better transportation (OR, 0.95; 95% CI, 0.92-0.98), and better sanitation (OR, 0.96; 95% CI, 0.93-0.98) were associated with reduced odds of depressive symptom. Conversely, greater population diversity (OR, 1.10; 95% CI, 1.04-1.16) were cross-sectionally associated with increased odds of depressive symptoms. Cox regression showed that people living in communities with higher population density (HR, 0.89; 95% CI, 0.86-0.92), higher education (HR, 0.95; 95% CI, 0.93-0.98), better economic conditions (HR, 0.94; 95% CI, 0.90-0.98) and better availability of social services (HR, 0. 96; 95% CI, 0.93-0.99) were likely to have lower risk of developing significant depressive symptoms over 7 years of follow-up, whereas population diversity (HR, 1.04; 95% CI, 1.01-1.07) slightly increased risk, after adjusting for covariates.

**Figure 1.**
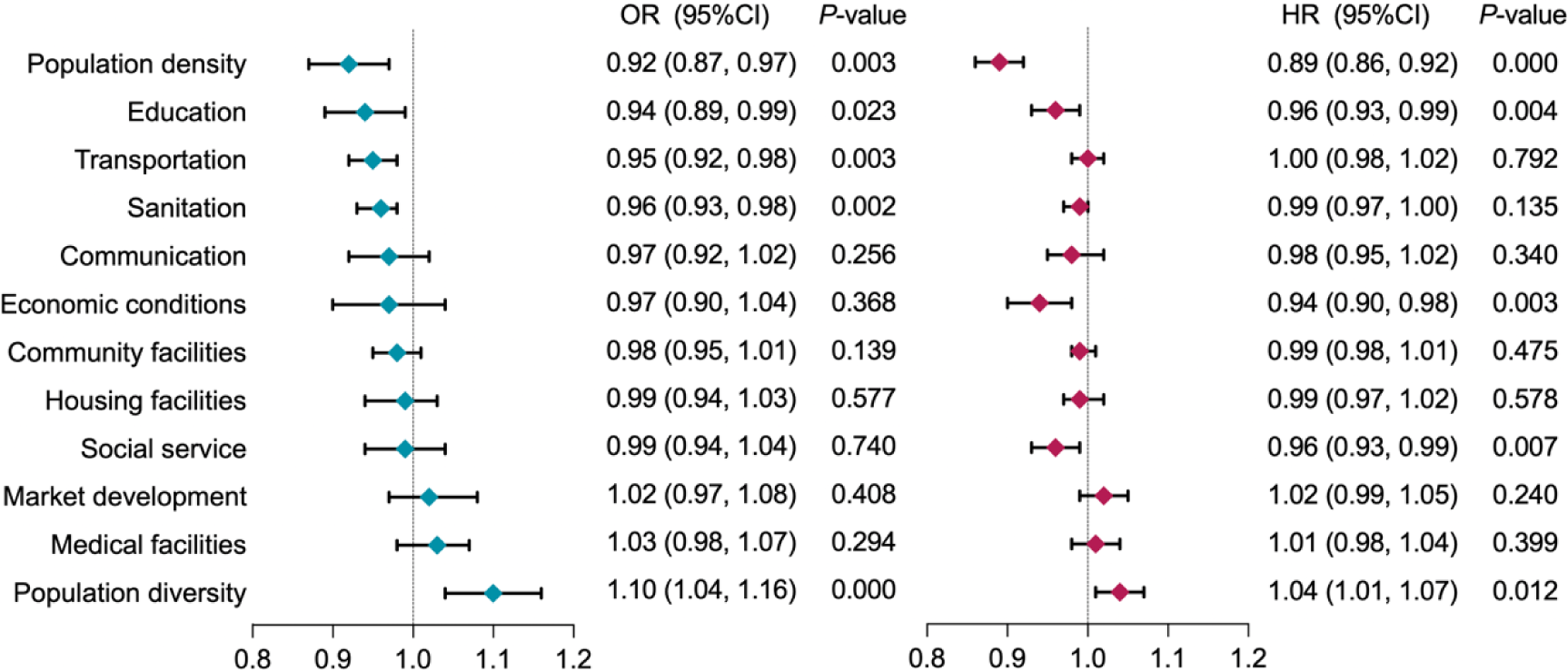
The cross-sectional and longitudinal associations between urbanicity components and significant depressive symptoms. Notes: All models adjusted for age, sex, education, marriage status, living alone status, smoking and drinking status, and chronic disease status. The urbanicity components are ordered by the odds ratios of the cross-sectional model. Abbreviations: OR, Odds ratio; HR, hazard ratios; 95% CI, 95% Confidence Intervals. OR were estimated by multilevel logistic models and HR by Cox regression models.

When stratified by residence or age group at baseline, urbanicity components display differential associations in urban and rural regions, as well as in middle-aged and older adults (Figure 2 and Table S11-S12). In urban settings, higher population density (OR, 0.93; 95% CI, 0.86-1.00) was associated with reduced, whereas higher population diversity (OR, 1.08; 95% CI, 1.00-1.17) was associated with increased depressive risks. In rural areas, higher population density (OR, 0.86; 95% CI, 0.81-0.92), higher education (OR, 0.92; 95% CI, 0.86 - 0.99), better transportation (OR, 0.95; 95% CI, 0.91-0.99) and better sanitation (OR, 0.94; 95% CI, 0.90-0.98) were associated with a reduced risk of depressive symptom, while higher population diversity (OR, 1.12; 95% CI, 1.04-1.20) was associated with increased risk. Among middle-aged group, higher population density (OR, 0.92; 95% CI, 0.86-0.98), better transportation (OR, 0.93; 95% CI, 0.89-0.96), better sanitation (OR, 0.95; 95% CI, 0.92-0.98) were associated with reduced, while higher population diversity (OR, 1.11; 95% CI, 1.05-1.18) were associated with increased depressive risks. For older adults, higher population density (OR, 0.91; 95% CI, 0.84-0.98) and better housing facilities (OR, 0.90; 95% CI, 0.84-0.97) were associated with reduced odds of depressive symptom.

**Figure 2.**
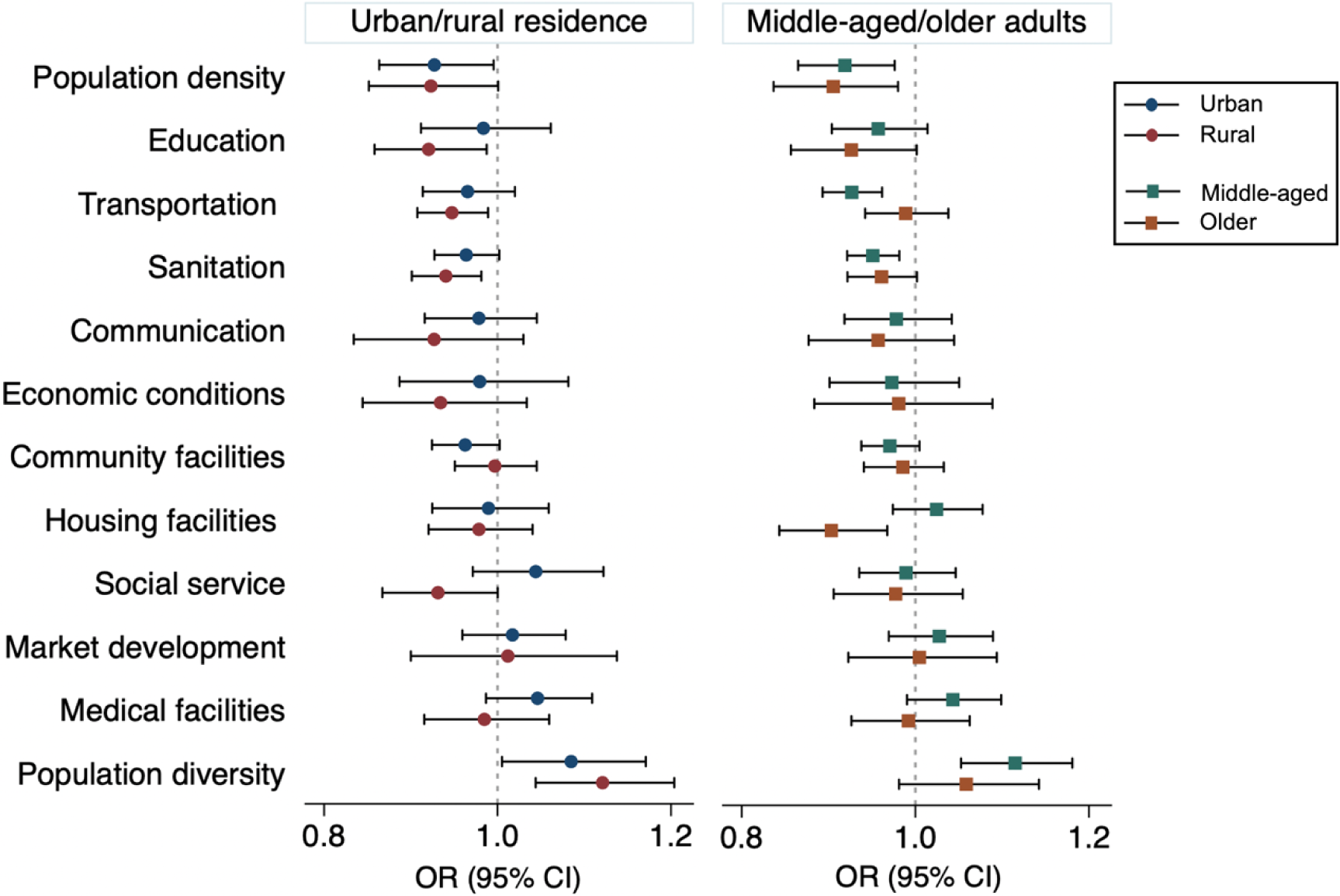
The cross-sectional associations between urbanicity components and significant depressive symptoms. Notes: Based on the baseline dataset, multilevel logistic models were used to estimate the effects of urbanicity components on significant depressive systems, stratified by residence and age group. All models were adjusted for age, sex, education, marriage status, living alone status, smoking and drinking status, and chronic disease status. The sample size of the total population was 12,515, with 4,875 urban residents, 7,640 rural residents, 9,218 middle-aged adults, and 3,297 older adults. The urbanicity components are ordered by the odds ratios of the cross-sectional model at baseline. The error bars represent the lower and upper boundaries of the 95% CI, with the line indicating the range of the OR value for each urbanicity component.

Figure 3 reports the risk of developing significant depressive symptoms by urbanicity components in longitudinal associations, stratified by residence and age group (Table S13-S14). In urban settings, higher population density (HR, 0.90; 95% CI, 0.86-0.95) and better social service (HR, 0.94; 95% CI, 0.90-0.99) were associated with reduced. Living in a rural area with higher population density (HR, 0.89; 95% CI, 0.85-0.93), better education (HR, 0.95; 95% CI, 0.91-0.98), communication (HR, 0.94; 95% CI, 0.89-1.00) and economic conditions (HR, 0.92; 95% CI, 0.87-0.97) and was associated with a lower likelihood of developing depressive symptoms, whereas residing in areas with higher population diversity (HR, 1.04; 95% CI, 1.00-1.08) was linked to an increased risk of depressive symptoms. For middle-aged adults, higher population density (HR, 0.90; 95% CI, 0.87-0.94), better education (HR, 0.95; 95% CI, 0.92-0.99), economic conditions (HR, 0.93; 95% CI, 0.89-0.98), and social service (HR, 0.96; 95% CI, 0.92-0.99) were associated with reduced, while higher population diversity (HR, 1.04; 95% CI, 1.00-1.07) was associated with increased depressive risks. For older adults, only population density (HR, 0.86; 95% CI, 0.80-0.92) had longitudinal association with developing depressive symptoms.

**Figure 3.**
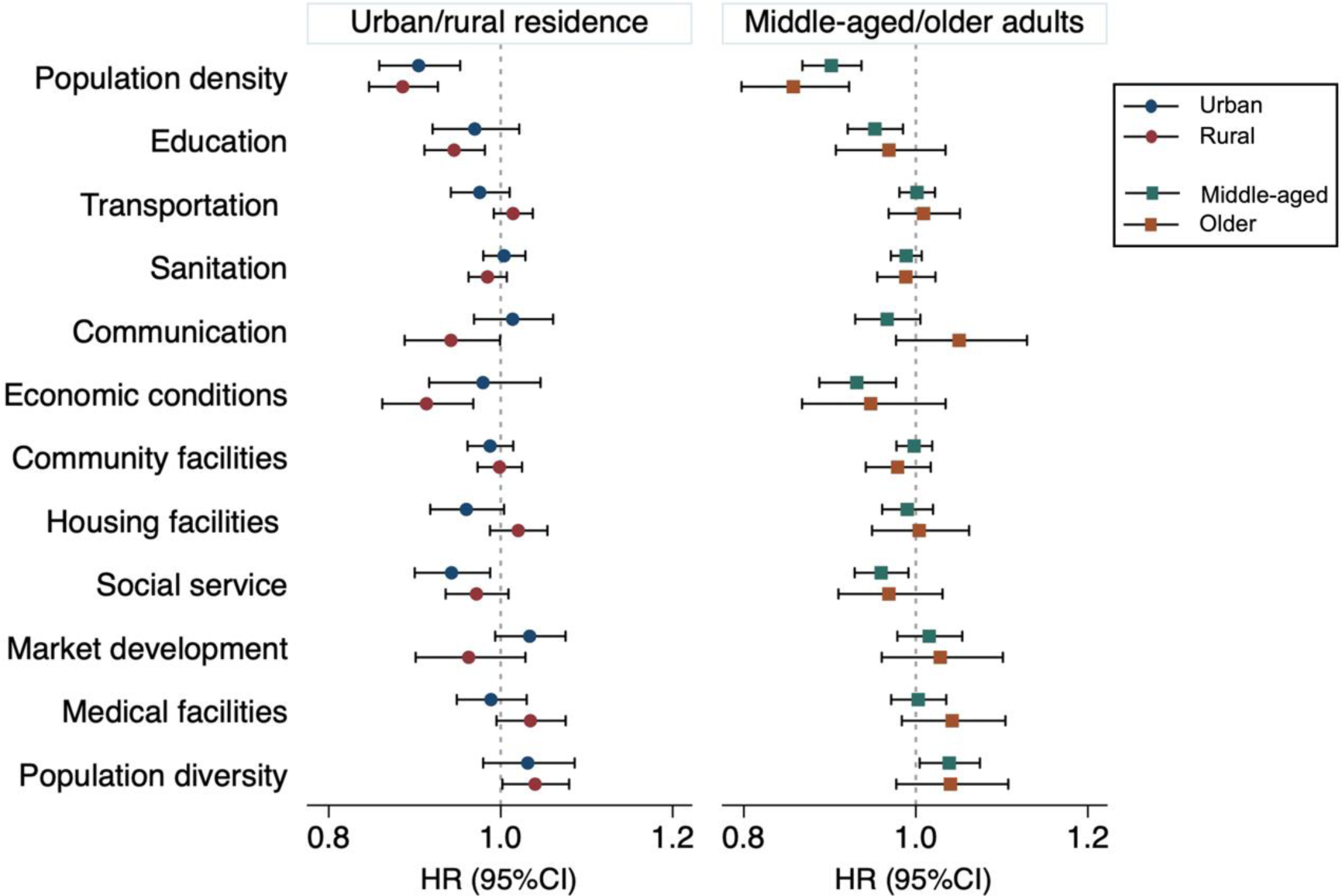
The longitudinal associations between urbanicity components and significant depressive symptoms. Notes: Based on the 7-years follow-up dataset, Cox regression models were used to estimate the effects of urbanicity components on significant depressive systems, stratified by residence and age group. All models were adjusted for age, sex, education, marriage status, living alone status, smoking and drinking status, and chronic disease status. The sample size of the total population was 8,766, with 3,558 urban residents, 5,208 rural residents, 6,733 middle-aged adults, and 2,033 older adults. The urbanicity components are ordered by the odds ratios of the cross-sectional model at baseline. The error bars represent the lower and upper boundaries of the 95% CI, with the line indicating the range of the HR value for each urbanicity component.

## Discussion

In this longitudinal study of middle-aged and older Chinese adults, we explored the associations between urbanicity and its components with significant depressive symptoms, in both cross-sectional and longitudinal associations. We found that a composite measure of urbanicity was associated higher risk of clinically-important depressive symptoms with the longitudinal associations being weaker than the cross-sectional ones. We examined the association of specific components of urbanicity, finding that higher population density, better transportation, sanitation, and education, along with lower population diversity, are cross-sectionally associated with a reduced risk of depressive symptoms. Better economic conditions and social services show additional effects in longitudinal associations. Further heterogeneity was found in the role of urbanicity components in affecting depressive symptoms between people living in urban and rural areas, and between midlife and older adults.

The observation that people living in areas with higher levels of urbanicity exhibit a higher risk of depressive symptoms diverges from previous research in HICs, where a higher degree of urbanicity has been associated with a greater prevalence of depressive disorder^16^, as seen in studies from the Netherlands^43^, Sweden^44^, and America^15^. Nonetheless, our results align with some other research conducted on the association between urbanicity and mental health within the Chinese context. Previous studies in China have typically used unidimensional urbanicity metrics such as the ratio of urban residents to the total population, population density, GDP per capita, and the proportion of secondary/tertiary industries to city-level GDP, and have indicated that depression is less prevalent in more urbanized areas^24,45^. Our study, using a more comprehensive urbanicity measurement, reinforces existing studies. The findings differ from those in HICs which reinforces that socio-cultural and economic contextual factors are likely to be crucial determinants of the effect of urbanicity on depression, and the need to examine which aspects of urbanicity are important in different contexts. Furthermore, this study extends beyond existing research by comparing cross-sectional and longitudinal associations. A weaker effect in the longitudinal association indicates that while urban living may initially provide mental health benefits, these advantages may diminish over time. It suggests that population-level urban planning strategies should consider temporal effectiveness, conduct dynamic monitoring of outcomes, and make timely adjustments.

Various population-related urbanicity components were found to be associated with the risk of depressive symptoms in both cross-sectional and longitudinal analysis. Our findings show that elderly individuals living in areas with higher population density have a reduced risk of depressive symptoms, which contradicts the findings of existing studies in high-income countries^46,47^. We extend previous findings from high-income nations regarding this association to a middle-income context, but we find an opposite result. Regarding population educational levels, another aspect of urbanicity, our findings are consistent with prior research^48^, showing that higher neighborhood educational attainment level was associated with lower depressive symptoms. We used income and education to calculate diversity index, representing the mean individual variation within an area and found that diversity was a risk factor for depressive symptoms, which confirms a prior study that has shown that neighborhood ethnic diversity is associated with mental health decline^49^. This may be because more people ‘unlike you’ in a diverse community may lead to worse mental health^50^, but the mechanisms of these links need further exploration. Based on these points, we may posit that social bonds and social segregation emerge as pivotal aspects that influence mental health in a society experiencing population growth during urbanization.

Longitudinally, we found that individuals living in areas with better economic conditions were associated with a decreased risk of depressive symptoms. This observation is consistent with a previous meta-analysis which suggested that poorer neighborhood socioeconomic conditions are associated with higher odds of depressive symptoms, and its potential mechanism is that people living in socioeconomically disadvantaged areas may experience greater stressors (e.g. exposure to violence, noise pollution) which have a detrimental impact on mental health^51^. China has developed a community-based service delivery model for social service. The elderly receive basic social welfare services from quasi-governmental community organizations in this model, providing important social support^52^. We furnished evidence for the longitudinal association between community-based social service and significant depressive symptoms over time, indicating that social service has a prolonged protective effect against developing depressive symptoms. These longitudinal associations might indicate that the beneficial effects of better economic conditions and social services on reducing the risk of depressive symptoms among older adults may take time to manifest. However, these improvements provide a solid foundation for gradually exerting positive effects.

Our findings confirm the cross-sectional associations between resources and infrastructure in neighboring areas and individual depressive symptoms, aligning with existing research^53,54^. Specifically, we found that improved transportation infrastructure and better sanitation facilities were associated with lower odds of depressive symptoms. People living in deprived areas may be more vulnerable due to increased stress in their lives and limited access to support^55^. For instance, the lack of sewage and waste management systems can contribute to water pollution. This not only leads to negative health consequences but also fosters negative emotions, such as insecurity and anxiety, due to limited access to clean water^56^. Moreover, accessibility to public transport broadens the range of activities available to individuals, providing older people with opportunities for a more active social lifestyle^57,58^.

Long-term economic reform in China has changed the population distribution between urban and rural areas, resulting in a geo-specific disparity in income, education, and healthcare^59^. This urban-rural gap may affect the mental health of older adults differently in urban and rural areas^60^. Our findings indicated that social service was associated with the risk of depressive symptoms among urban residents, whereas components such as economic conditions, transportation, communication, sanitation, and education were significantly associated with their rural counterparts. These results underscore the importance of tailoring intervention strategies to address urban environmental determinants of mental health according to the specific urban or rural context. It also indicates that it is not merely urbanicity or rurality per se but the specific differences in living conditions that matter. Our findings support prior research that rural community infrastructure improvement is vital for minimizing the mental health gap between rural and urban people^61^. In addition, this study revealed that urbanicity has a greater effect on rural participants compared to their urban counterparts. As China’s ongoing rural revitalization strategy advances, involving improvements in water supply, infrastructure, and public education^62^, it is expected that the rural-urban gap of mental health will narrow in the future.

When designing population-based strategies to prevent depressive symptoms, a life course perspective is essential, as modifiable risk factors in midlife differ from those in later life^63^. We stratified and compared the effects between middle-aged and older adults and found that more urbanicity elements in this study impact the risk of depressive symptoms among middle-aged adults. For older adults, the two urbanicity components significantly associated with lower depressive symptoms are higher population density and better housing facilities. This may be explained by the fact that higher population density might help to reduce social isolation by providing more opportunities for social interaction and support^64^. In addition, better housing facilities is likely to provide a comfortable and safe living environment, thereby contributing to their mental health^65^. In comparison, middle-aged adults are more active, mobile, and engaged in the work and social life, making urbanicity aspects such as transport, sanitation, social services, education, and economic conditions more critical to their mental health and well-being. The need for age-specific urban planning and public health interventions is highlighted by these age differences.

## Limitation

The present study had several limitations. Firstly, although the CES-D is a commonly used tool for measuring depressive symptoms, it relies on self-report and inevitably has potential information bias. Second, community data was only collected at the baseline survey, which means our study could not account for the changes of urbanicity characteristics between survey waves or track urbanization developments over time. Future research with more waves of data may provide a more comprehensive understanding of the impact of urbanicity. Additionally, the community questionnaire was completed by village or community officials. Thirdly, future studies should also consider other important environmental factors that accompany the urbanization process, such as pollution. Fourth, despite these findings providing valuable insights, further research into the underlying causes and mechanisms of the effects of these urbanicity components is required before they can be used to inform policy and public health interventions. Finally, the direction of causation of the associations is unclear as people with worse mental health may be more likely to move to areas with adverse social conditions, as per the social drift hypothesis^66^.

## Conclusions

This study advances our understanding of the association between urbanicity and depressive symptoms in middle-aged and older adults. To the best of our knowledge, this is the first study of its kind, and the innovative findings offer new insights into depression in relation to urban environmental factors. Our findings underscore the multifaceted impact of urban living on risk of developing depressive symptoms, by using a multidimensional urbanicity scale to delve deeper into the complex urbanicity-mental health nexus. The varying associated urbanicity components among subgroups suggest that the real impact might originate from the characteristics of their urban life. Future research should consider the impact of these components of urbanicity in other settings and populations. It identifies some important urbanicity components that can decrease the risk of depressive symptoms and paves the way for future research to investigate the mechanisms underlying these linkages. Taking into account the time effectiveness of urban planning, the resource constraints in rural areas and the different daily life characteristics of older adults, tailored policies hold greater potential to reduce the burden of depressive symptoms. Our findings may contribute to public health policy and guidelines and novel population-based interventions aimed at reducing depression in community settings.

## Data Availability

All data produced are available online at the China Health and Retirement Longitudinal Study (CHARLS, accessible at http://charls.pku.edu.cn/).

